# Targeting a hypothetical estimand based on adherence with the parametric g-formula: A post hoc secondary analysis of the MET-PREVENT randomised controlled trial

**DOI:** 10.1101/2025.07.03.25330760

**Authors:** Shaun Hiu, Nina Wilson, Kevin Wilson, Nan Lin, Miles D Witham, James M S Wason, The MET-PREVENT Study Group

**Affiliations:** Population Health Sciences Institute, Newcastle University, Newcastle upon Tyne, UK; School of Mathematics, Statistics and Physics, Newcastle University, Newcastle upon Tyne, UK; AGE Research Group, Translational and Clinical Research Institute, Faculty of Medical Sciences, Newcastle University, Newcastle upon Tyne, UK; NIHR Newcastle Biomedical Research Centre, Newcastle upon Tyne Hospitals NHS Foundation Trust, Cumbria, Northumberland, Tyne and Wear NHS Foundation Trust and Newcastle University, Newcastle upon Tyne, UK; Nuffield Department of Primary Care Health Sciences, University of Oxford, Oxford, UK

**Keywords:** Estimands, Causal inference, G-formula, Mediation, Secondary analysis, Quantitative bias analysis

## Abstract

**Background:** We conducted a post hoc secondary analysis of the MET-PREVENT randomised placebo-controlled trial to target a hypothetical estimand – the mean difference in physical performance, as measured by the 4-metre walk speed at four months after randomisation, between treatment arms – under the hypothetical scenario that all participants adhered to assigned treatment.

**Methods:** We viewed the targeting of this hypothetical estimand as a type of causal mediation problem, where randomisation is a binary exposure, adherence is a binary mediator, there is an exposure-mediator interaction, and mediator-outcome confounders that are caused by the exposure (e.g., gastrointestinal symptoms) are present. We used the parametric g-formula to estimate the average controlled direct effect (CDE) of metformin (versus placebo) on 4-metre walk speed, which is interpreted as the average treatment effect under the hypothetical scenario that all trial participants adhered to assigned treatment. Variables identified as confounders were informed by a literature review and discussions with an expert; assumptions about the causal structure were represented in a directed acyclic graph. We applied a probabilistic bias analysis (PBA) to understand the potential for bias assuming adherence had been misclassified; observed adherence based on returned tablet count may be an inaccurate version of “true adherence” based on actual consumption.

**Results:** Our sample size was 70 trial participants (34 metformin, 36 placebo). Our estimate of the CDE was 0.072 [percentile-based bootstrap 95% CI −0.292, 0.445]. Results from PBA indicated that the greater the extent of misclassification, the more the CDE may be estimated with bias and with over-optimistic precision.

**Conclusions:** Our study provided supporting information on metformin’s potential role as a repurposed medication to improve physical performance in nondiabetic older adult patients with physical prefrailty/frailty and probable sarcopenia. Unlike the main trial results, our results do not rule out the possibility of a meaningful benefit of metformin, provided that full adherence can be assured. We highlighted the parametric g-formula as a useful method in trials to target hypothetical estimands that are concerned with treatment adherence.

**Trial registration:** ISRCTN29932357

## Introduction

The de facto position of randomised placebo-controlled superiority trials is to make conclusions from effects that preserve the intention to treat (ITT) principle, which is analogous to the treatment policy estimand, followed by accompanying per-protocol analyses whereby non-adherers are either excluded or censored. It is known that such ITT effects are particularly sensitive to intercurrent events since any event occurring after randomisation is essentially ignored in the analysis (1). Treatment adherence may be one such intercurrent event, and differential adherence rates between arms (whereby the intervention arm usually has the lower adherence rate possibly due to the adverse effects of the new treatment) make it challenging to separate between two propositions for observed null findings from ITT results: either the treatment regime under investigation is inefficacious in the target population or it is efficacious but was observed to be inefficacious due to low adherence. Making this distinction is important because adherence is a behaviour that can be improved through intervention.

In this study, we conducted a post hoc secondary analysis of the MET-PREVENT trial to target a hypothetical estimand with the parametric g-formula to estimate the average controlled direct effect (CDE) of metformin (versus placebo) on 4-metre walk speed under the hypothetical scenario that all participants adhered (2).

We viewed the targeting of this hypothetical estimand as a type of causal mediation problem, where randomisation is a binary exposure, adherence is a binary mediator, there is an exposure-mediator interaction (because adhering to metformin may matter more than adhering to placebo), and presence of mediator-outcome confounders that are caused by the exposure (e.g., gastrointestinal symptoms). The parametric g-formula (also known as g-computation) overcomes limitations of conventional methods in investigating causal effects when adherence plays the role of a mediator. Traditional per-protocol analyses that involve restricting the sample to those who are adherent (effectively censoring non-adherers) are at-risk of selection bias as an event that happens after randomisation is conditioned upon. Valid estimation of per-protocol (PP) effects requires controlling for factors that influence adherence (i.e. mediator-outcome confounders) (3, 4). However, where there is exposure-mediator interaction and mediator-outcome confounders are caused by the exposure, there is a need to turn to a family of methods known as “g-methods” of which the g-formula is a member (5).

## Methods

### Details of the trial

MET-PREVENT was a multicentre, proof-of-concept, superiority randomised controlled trial (RCT) of older adults with probable sarcopenia and frailty or prefrailty, but without diabetes mellitus, to investigate if metformin, a glucose-lowering therapy, was efficacious in improving physical performance in this population. The primary analysis was an intention-to-treat (ITT) analysis which found no statistically significant difference between arms (0.001 [95% CI −0.06, 0.06] m/s). The primary results have recently been accepted for publication (6). Adherence to medication was observed only at the end of the trial when tablets were returned and counted; a participant was defined as adherent if they consumed ≥80% of the expected number of tablets they should have taken by the final four-month visit. There was a noticeable difference in adherence rates between the intervention (53%) and placebo (78%) arms respectively.

Seventy-two participants were randomised 1:1 (using minimisation with a 30% random element) between metformin and matched placebo. The primary outcome was the 4-metre walk speed test at a four month visit (7). A minimal clinically important difference (MCID) of 0.1 m/s was pre-specified. One participant withdrew before receiving their medication and one died during follow-up, leaving n=70 with available data on the primary outcome (4-metre walk speed). Because there was data on only one death and study withdrawal respectively, we could not model these events. Scheduled visits occurred at baseline, one month, two months, and three months after randomisation, with a final visit at four months where the primary outcome was measured. No participant was lost to follow-up. Details on the trial eligibility criteria and design are available in a published protocol (7). The trial is registered on ISRCTN (ISRCTN29932357).

Treatment assignment is regarded as the exposure in our secondary analysis. Participants were randomised to receive metformin 500mg or matching placebo tablets. Dosage was three tablets per day. All participants were supplied with 372 tablets at the point of medication reception to last the entire four-month follow-up duration.

True adherence, based on knowledge of actual tablet consumption, was regarded as the mediator. However, in the absence of this knowledge, observed adherence, defined as a medication possession ratio (MPR) ≥ 80% at the final visit when unused tablets were returned and counted, was treated as the closest (but potentially misclassified) surrogate to true adherence in the data. The MPR could not be computed for two participants in the intervention arm as tablets were disposed of by accident. One participant was classified as non-adherent as they experienced gastrointestinal symptoms leading to treatment discontinuation at five days after randomisation. The other participant was classified as adherent based on mean imputation of the MPR.

The primary outcome was the 4-metre walk speed (in m/s) measured at the four-month visit. All participants were tasked to walk 4-metres on a measured course and at a pace they normally walk at if they were walking down the street. Participants were allowed to use their walking aid. The faster of two attempts was the primary outcome.

### Hypothetical estimand

Our estimand of interest is the mean difference in physical performance, as measured by the 4-metre walk speed at four months after randomisation between metformin and placebo arms – under the hypothetical scenario that all participants adhered to assigned treatment over a four-month period, amongst physically pre-frail and frail nondiabetic older adults with probable sarcopenia.

### Assumptions about underlying causal relationships

We make explicit our assumptions about the underlying causal relationships in a directed acyclic graph (DAG) (Figure 1); we did not assume unmeasured exposure-outcome confounding owing to randomisation. Variables identified as mediator-outcome confounders were informed by the intersection between literature on factors which influence adherence to medication (8–11) and predictors of walk speed amongst older adults, and discussions with an expert (MDW).

**Figure 1:**
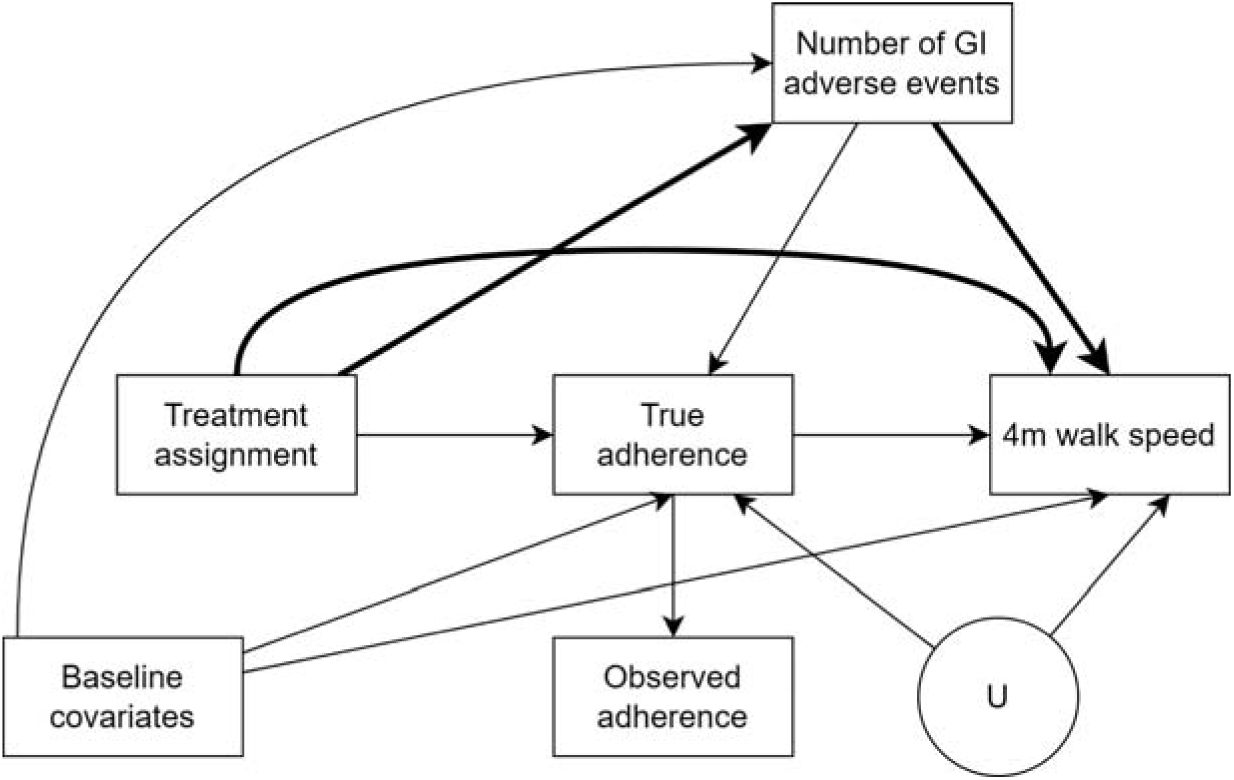
Directed acyclic graph representing assumptions of causal relationships. The observed adherence variable in our data (which is based on returned tablet counts) may be a misclassified version of “true adherence” (which is based on actual consumption and not known to us). The variable U represents unmeasured mediator-outcome confounder(s) that are not available in the data. The bolded paths collectively make up the CDE of treatment assignment on the outcome.

Baseline mediator-outcome confounders included age (12, 13), sex (12), obesity (body mass index ≥30 kg/m^2^) (14, 15), history of depression (16, 17), functional ability as defined by the Nottingham Extended Activities of Daily Living (NEADL) (13), polypharmacy (≥5 concomitant medications) (18), and baseline 4-metre walk speed. We postulated that individuals who were more impaired on their walk speed may be more motivated to adhere given that participants were informed of the premise of the trial and blinded to treatment allocation. The mediator-outcome confounder caused by the exposure was the number of gastrointestinal adverse events over follow-up (0, 1, or ≥2 events). Metformin is known to potentially cause gastrointestinal (GI) symptoms, such as diarrhoea and nausea, which may discourage adherence (8). We postulated two ways in which GI symptoms may influence walk speed. Participants who experience GI symptoms may perform worse on physical performance measures, especially if the GI side effects made them less active and they became physically deconditioned. Alternatively, if metformin’s effect on skeletal muscle is due to a change in the gut microbiome, it may be that only those who do experience GI symptoms achieve benefit. Adverse events were recorded in an adverse events log and GI adverse events are defined as those which were coded under “gastrointestinal disorders” within the Medical Dictionary for Regulatory Activities (MedDRA®) terminology (version 25) (19). We replaced the missing subscale items on the NEADL with the participant-specific median from the completed items in the subscale, provided less than three items within a subscale were missing.

### Statistical analysis

We used the parametric g-formula to identify the CDE. We refer readers to Appendix A where we introduce the notation for potential outcomes and introduce the g-formula. A stepwise process for the parametric g-formula and our modelling assumptions are described in Appendix B. We reported the estimate of the CDE and a percentile-based 95% confidence interval (CI). We also reported a bias-corrected and accelerated (BCa) bootstrap 95% CI for comparison; the relevance of the BCa bootstrap is included in our discussion but, briefly, it has been proposed to improve the coverage of the parametric g-formula estimator, especially for small sample sizes (20). The g-formula algorithm is provided in greater detail elsewhere (21). Analyses were carried out in R version 4.3.0.

### Sensitivity analyses

We assessed how well our model specifications agreed with our assumptions about the underlying causal structure by estimating the natural course (22). The idea of the natural course is if one were to derive the observed sample mean of the primary outcome through the parametric g-formula, it would require us to specify parametric models for the conditional probability of treatment, adherence, occurrence of GI adverse events, and the conditional mean of the primary outcome. If all these models were correctly specified, then the summary statistics and distributions of the variables that are predicted by the parametric g-formula algorithm should approximately match the observed data. We compared the mean, standard deviation, and empirical cumulative distribution function (eCDF) of the observed primary outcome data against the data predicted by the parametric g-formula, and the means of observed adherence and GI adverse events data against the those predicted by the parametric g-formula.

Observed adherence rates based on returned tablet count may overestimate “true adherence” rates based on actual consumption, and there may be concerns over bias due to a misclassified mediator. To understand the impact of misclassifying adherence, we applied a probabilistic bias analysis (PBA) that adjusted the ‘naïve’ estimate of the CDE for misclassification by incorporating information from the literature surrounding the possible extent of misclassification (23). The aim of a PBA was to gain insight into the impact that misclassification had on the magnitude and direction of bias in our ‘naïve’ CDE estimate, as well as on our uncertainty (24). We simulated four conditions that represented: 1) a very high certainty in our beliefs that there is a low degree of misclassification; 2) a very high certainty in our beliefs that there is a severe degree of misclassification; 3) more moderate certainty in our beliefs that there is a low degree of misclassification; and lastly 4) more moderate certainty in our beliefs that there is a severe degree of misclassification. Detailed methodology on the PBA is available in Appendix C. We reported the median and percentile-based 95% simulation interval (95% SI) of the misclassification bias-adjusted CDE. The misclassification bias-adjusted point estimate was compared against the naïve point estimate, as well as the widths of the simulation intervals versus the confidence interval (24).

## Results

Summary characteristics of our sample are in Table 1. Thirteen participants in the control arm (36.1%) and 27 participants in the intervention arm (79.4%) experienced at least one GI adverse event over the course of follow-up. Of the 27 in the control arm, 10 discontinued due to GI adverse events (9 participant-led, 1 investigator-led). Of the 13 in the control arm, 1 discontinued due to a GI adverse event which was participant-led. Two participants assigned to metformin discontinued due to elevated lactate concentrations (>4 mmol/L at the one-month and three-month visits respectively) and one participant assigned to placebo discontinued due to hypoglycaemia (<4 mmol/L) at the two-month visit. None discontinued due to severe renal impairment.

**Table 1:** Characteristics of the sample.

|  | Analysis set (n=70) |
| --- | --- |
| Treatment arm, n (%) |  |
| Metformin | 34 (48.57) |
| Placebo | 36 (51.43) |
| Age, years, mean (SD) | 80.49 (5.75) |
| Sex, n (%) |  |
| Female | 41 (58.57) |
| Male | 29 (41.43) |
| History of depression, n (%) | 19 (27.14) |
| Obesity (body mass index $\geq 30$ kg/m<sup>2</sup>), n (%) | 18 (25.71) |
| Nottingham Extended Activities of Daily Living, mean (SD) | 15.53 (4.94) |
| Polypharmacy ($\geq 5$ medications), n (%) | 60 (85.71) |
| Baseline 4-metre walk speed, m/s, mean (SD) | 0.59 (0.22) |
| Four month 4-metre walk speed, m/s, mean (SD) | 0.58 (0.22) |
| Observed adherence, n (%) | 47 (67.14) |
| Number of gastrointestinal adverse events over follow-up, n (%) |  |
| 0 | 30 (42.86) |
| 1 | 30 (42.86) |
| 2 | 2 (5.71) |
| 3 | 3 (7.14) |
| 4 | 4 (1.43) |

Our estimate of the CDE was 0.072 [percentile-based bootstrap 95% CI −0.292, 0.445] m/s. We also reported the BCa bootstrap 95% CI [-0.244, 0.519] for comparison. We conducted a natural course analysis to validate our models used in the parametric g-formula. We observed agreement, to an extent, between the observed data and the data predicted from the parametric g-formula algorithm under the natural course (Table 2). The eCDFs of the observed and predicted primary outcome data are presented in Figure 2. We observed that there was close agreement on the proportion of GI adverse events, adherence, and the distribution of the primary outcome. Our PBA results are presented in Table 3.

**Figure 2:**
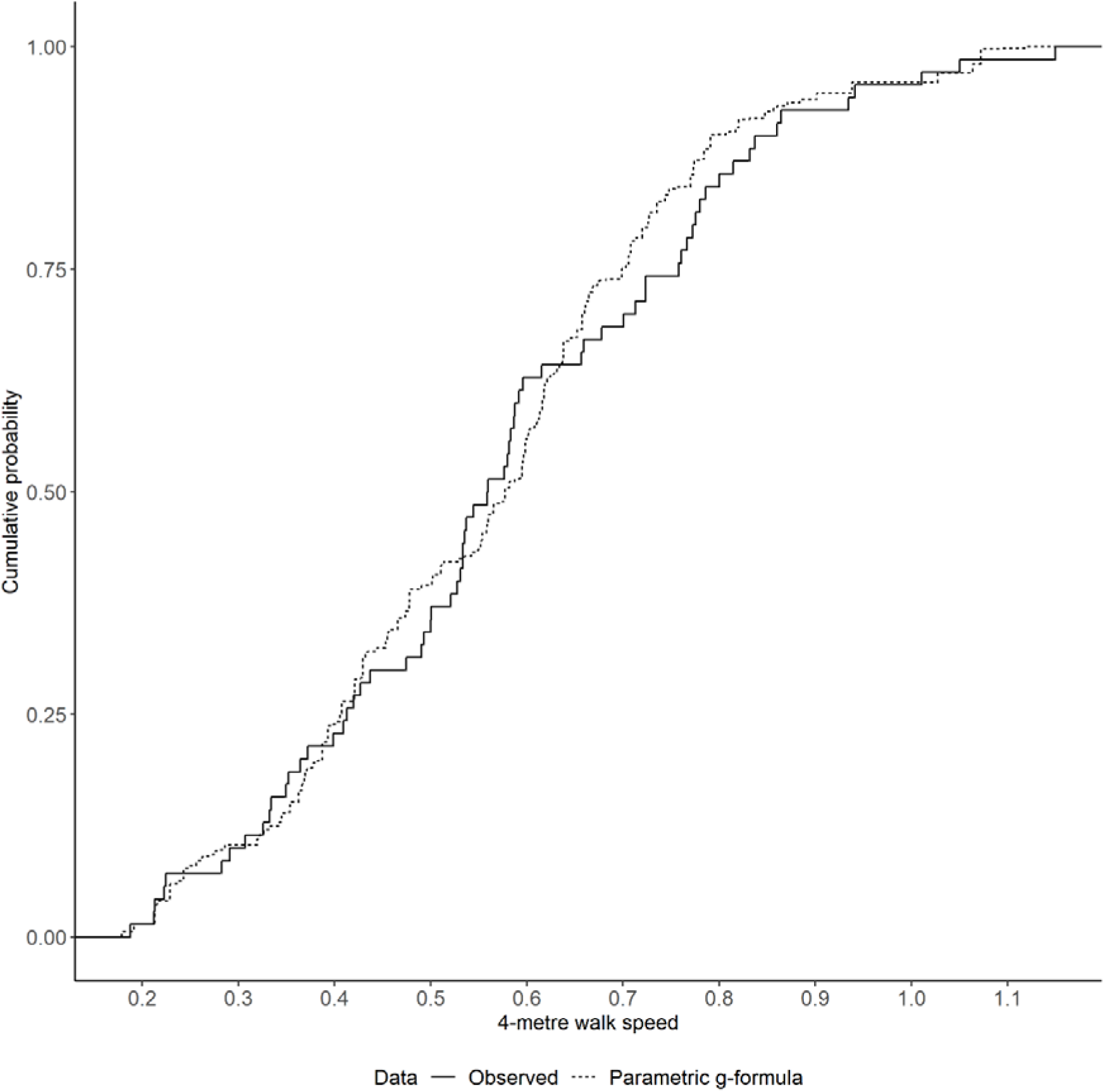
Empirical cumulative distribution functions of the observed and predicted primary outcome data.

**Table 2:** Comparison of observed data and data predicted by the parametric g-formula estimator in the natural course analysis.

|  | Observed data (n=70) | Predicted data (n=10000) |
| --- | --- | --- |
| **Number of GI adverse events** |  |  |
| No events, % | 42.86% | 40.66% |
| One event, % | 42.86% | 45.04% |
| ≥2 events, % | 14.29% | 14.30% |
| **Adherence** |  |  |
| Observed adherence, % | 67.14% | 66.80% |
| **4-metre walk speed at four months, m/s** |  |  |
| Mean (SD) | 0.58 (0.22) | 0.56 (0.20) |
| 1<sup>st</sup> percentile | 0.20 | 0.19 |
| 2.5<sup>th</sup> percentile | 0.21 | 0.21 |
| 5<sup>th</sup> percentile | 0.22 | 0.23 |
| 25<sup>th</sup> percentile | 0.41 | 0.41 |
| 50<sup>th</sup> percentile | 0.56 | 0.58 |
| 75<sup>th</sup> percentile | 0.75 | 0.70 |
| 95<sup>th</sup> percentile | 0.94 | 0.94 |
| 97.5<sup>th</sup> percentile | 1.02 | 1.06 |
| 99<sup>th</sup> percentile | 1.08 | 1.07 |

GI, gastrointestinal; SD, standard deviation.

**Table 3:** Results of probabilistic bias analysis.

| Scenario | Description | Prior Beta distribution of the FPR | Median [95% SI] |
| --- | --- | --- | --- |
| 1 | Very high certainty of low degree of misclassification | Beta(33.3, 231) | 0.066 [-0.657, 0.818] |
| 2 | Very high certainty of severe degree of misclassification | Beta(355, 205) | 0.020 [-0.744, 0.760] |
| 3 | Moderate certainty of low degree of misclassification | Beta(4, 26) | 0.069 [-0.687, 0.809] |
| 4 | Moderate certainty of low degree of misclassification | Beta(39.8, 25) | 0.029 [-0.728, 0.779] |

FPR, false positive rate; SI, simulation interval.

## Discussion

Our post hoc secondary analysis supports the trial’s primary finding of insufficient evidence to conclude a beneficial effect of metformin on older adults with physical pre-frailty/frailty and probably sarcopenia. However, in contrast to the prespecified primary analysis findings, our results do not rule out the possibility of either a meaningful benefit of metformin, provided that the consumption of ≥80% of prescribed tablets can be assured. Both the percentile-based and BCa bootstrap CIs of our CDE included zero as well as the MCID on both sides of the null, indicating that a null effect, meaningful benefit, and meaningful harm are all consistent with our analysis (25). Our main analyses assumed that there was no misclassification of treatment adherence and that observed adherence faithfully reflected true adherence. If on the other hand we suspect there to be misclassification, our sensitivity analyses indicated that the greater the extent of misclassification, the more the CDE may be estimated with bias (estimate would have been closer to the null if true adherence had been collected and used in the parametric g-formula analysis) and with over-optimistic precision (confidence intervals would have been wider if true adherence had been collected and used in the parametric g-formula analysis). Because the actual extent of misclassification is not known, we ran our PBA under multiple scenarios to cover different sets of beliefs about the misclassification. It could be argued that the low misclassification scenarios were more plausible than the severe misclassification scenarios. This is based on the premise that participants on clinical trials are generally motivated to follow trial processes especially if an investigational product has the potential to improve the symptoms of a progressive disorder like sarcopenia. Hence, the plausible source of misclassification may be benign forgetfulness such as accidental disposal as opposed to an active intent to defy protocol.

### Strengths

Methods based on the potential outcome framework have utility in overcoming some of the limitations of traditional per-protocol methods which may unintentionally subject themselves to bias (4). To illustrate this, we compare our estimates with the those from the trial’s original PP analysis of 0.023 [95% CI −0.053, 0.099] m/s. The noticeable difference between the point estimates may reflect that the original PP analysis was subjected to collider-stratification bias when adherence was conditioned upon (by limiting the analysis to adherers only) (26, 27). A collider is variable *C* that is caused by two other variables *A* and *B* (hence the effects of the two variables on *C* ‘collide’). The effect of A on B is estimated with (collider-stratification) bias when a model regressing *B* on *A* also includes *C* as a covariate. With reference to Figure 1, and assuming that it accurately reflects the underlying causal structure, limiting the analysis to adherers only induces a noncausal association between treatment assignment and the outcome through the baseline covariates and number of GI adverse events because adherence is a collider on four backdoor paths through GI adverse events. If one were to adjust for the baseline covariates and number of GI adverse events in a traditional outcome regression model (e.g., analysis of covariance), a portion of the direct effect of treatment assignment on the outcome which goes through number of GI events would be removed. Hence, the need for g-methods to avoid collider-stratification bias, unintended partial subtraction of direct effects, and importantly interpret results as causal effects. A more detailed introduction to directed acyclic graphs, collider-stratification bias, and backdoor paths is available elsewhere (27).

The strength of the parametric g-formula is that parametric models allow investigators to extrapolate beyond the observed data, thus positivity violations due to sparsity of data are less of an issue compared to methods using inverse probability weights (28). However, this trade-off means the parametric g-formula relies heavily on correct model specification to extrapolate over strata of covariates with sparse data. We attempted to make appropriate model specifications as much as possible by referencing prior literature and seeking expert opinion on potential mediator-outcome confounders, and relaxing modelling assumptions by allowing for interactions between treatment and all covariates and the inclusion of non-linear terms of continuous covariates in the primary outcome model. On the other hand, baseline cognitive functioning (8, 10, 29, 30) may be an unmeasured mediator-outcome confounder such that higher cognitive functioning may lead to better adherence and walk speed. Under unmeasured confounding, a qualified interpretation of our estimates is necessary. If the confounder(s) exists and it increases the likelihood of adherence and the average value of the outcome simultaneously, then our estimate of the CDE may be interpreted as estimates for the lower bound of the true CDE instead (2). The impact of this confounder was partially mitigated against as none of the participants was diagnosed with dementia and that baseline walk speed and functional ability are correlated with cognition (31, 32). Nevertheless, this means that our estimate of 0.072 m/s may be regarded as our best estimate of this lower bound, and the true effect of metformin under unanimous adherence may be larger than this quantity. However, given the confidence interval, a negative-valued lower bound is consistent with our data, which maintains the possibility of the true CDE being zero.

### Limitations

The use of the parametric g-formula at small sample sizes carries limitations. Few studies have evaluated the performance of the parametric g-formula estimator in small sample sizes (n ≤100). Tackney and colleagues (20) examined the performance of the parametric g-formula estimator of the average treatment effect (ATE) of a point exposure on a continuous outcome. They examined the performance of the estimator when sample size was small (n=50 and n=100) and the model for the conditional mean of their outcome included a treatment variable, 17 covariates (which were all predictive of the outcome), and interactions between treatment and all covariates. Provided all confounders have been accounted for, their results indicate that whilst the estimator had a mean bias close to zero, but there was suboptimal coverage (approximately 90% at either sample size) which were due to the model-based standard errors underestimating the empirical standard error. Coverage was further reduced when including four additional covariates that were unpredictive of the outcome. These findings regarding suboptimal coverage are supported by Chatton and colleagues (33) who evaluated the performance of the parametric g-formula estimator of the average treatment effect of a point exposure when the outcome was binary, sample size was 100, and the outcome model included 9 covariates (6 were predictive of the outcome) and the treatment variable. Assuming correct model specification, their results indicated that the estimator had a mean bias close to zero but coverage was suboptimal at 91.8%; reducing the number of covariates appeared to shift the coverage probability closer towards the desired 95% probability. Small sample corrections such as BCa bootstrap have thus been proposed to improve coverage (20).

A limitation of the CDE is that all participants are always set to adhere through some conceivable intervention. This may not be realistic as interventions should allow treatment to be discontinued at any point when a participant experiences a clinical event that jeopardises their safety. Relevant events in the context of metformin would be elevated lactate concentrations, hypoglycaemia, and severe renal impairment. One strategy would have been to conceive of a dynamic intervention on adherence that allows for non-adherence when participant safety is compromised at any point during follow-up (34). Given that the incidence rates of elevated lactate concentrations, hypoglycaemia, and severe renal impairment following metformin use are low based from previous reports and our observations (35–37), it may be reasonable to interpret our effect as a close approximation to the effect under this dynamic intervention on adherence.

We observed a relatively wider confidence interval of the CDE compared that of the trial’s ITT and PP effects. One possible reason could be that controlled direct effects estimated by the parametric g-formula have greater sample size requirements for the same level of precision because of the added modelling assumptions. A previous study conducted a secondary analysis of a placebo-controlled trial to investigate the per-protocol effect of complete adherence to low-dose aspirin at preconception to improve live births amongst 1,227 women with documented pregnancy losses and were actively trying to conceive (38). Compared to the trial’s ITT effect of a relative risk of live birth of 1.10 [95% CI 0.98, 1.22], the parametric g-formula estimate of the per-protocol effect in which all participants adhered was 1.33 [95% CI 1.08, 1.64]; an approximately two-fold increase in the width of the confidence interval on the log relative risk scale. Another reason could be that at the given sample size the models were overfitted to the data. However, Tackney and colleagues (20) showed by simulation that even at a fixed sample size of 50, increasing the number of covariates or even adding interaction terms between treatment all covariates did not substantially change the model-based standard error or empirical standard error of the parametric g-formula estimator of the ATE of a point exposure on a continuous outcome. Further work could be developed to investigate if this also extends to mediation analyses.

### Implications for trial methodologists

Our study allowed us to draw out lessons for trialists. We highlighted the parametric g-formula as a useful method in trials to target hypothetical estimands that are concerned with treatment adherence. Our study shines a light on two contexts in which targeting a hypothetical estimand based on adherence is meaningful.

Firstly, efficacy trials may benefit most whereby maximising adherence is important so as not to miss a possible benefit of treatment. Meeting the threshold for adherence for all participants in an efficacy trial setting could possibly be reached perhaps by introducing an initial titration period (39) plus raising awareness amongst participants about the nocebo effect i.e. a portion of GI adverse events may be attributable to negative expectations about the medicine rather than a pharmacological effect (40). Other strategies may also include patient education and counselling (8), shared decision-making between clinicians and patients (8), dose adjustment (41), use of extended release formulations (41), and lifestyle adaptations (e.g., time medication intake around meals (41)). However, in effectiveness trials, it is much more important that the trial conditions reflect how the drug will be used in the real world, so if low adherence is going to be an issue in clinical practice, seeing this in the trial is preferable and thus there may not be much to be gained from targeting an adherence-based hypothetical estimand.

Secondly, while there may be no substitute for efforts to actually maximise adherence at the design and implementation stages, methods such as the parametric g-formula to target adherence-based hypothetical estimands may be the best alternative when maximising adherence is challenging in practice. If there is an a priori interest in the treatment effect under an ideal circumstance of unanimous adherence, then there may be a need to build in an analysis to estimate the CDE at the trial design stage as it may influence sample size. Generally, closed form solutions are not readily available for causal effects thus Monte Carlo simulation will be needed (42, 43).

## Ethical standards

All participants provided their informed consent. The trial was approved by the UK Health Research Authority North-West - Liverpool Central Research Ethics Committee (20/NW/0470) and the UK Medicines and Healthcare products Regulatory Agency (2020-004023-16). The trial was sponsored by the Newcastle Upon Tyne Hospitals NHS Foundation Trust. The trial complied with the ethical standards laid down in the 1964 Declaration of Helsinki and its later amendments.

## Data statement

Data can be shared upon reasonable request to the NIHR Newcastle Biomedical Research Centre by contacting Professor Miles Witham (Chief Investigator).

