## Supplemental material for "Targeting a hypothetical estimand based on adherence with the parametric g-formula: A post hoc secondary analysis of the MET-PREVENT randomised controlled trial"

**Appendix**

### **Appendix A: Introduction to potential outcomes and the g-formula**

*The g-formula for point exposure*

In this appendix, we briefly describe potential outcomes, the g-formula, and the parametric g-formula. The g-formula is a generalisation of the method of standardisation to identify the marginal effects from the observed data (1). A potential outcome, such as $Y^{a}$, is a quantity that represents the value an outcome $Y$ would take *if hypothetically* the individual received a particular treatment strategy. For example, initiate treatment $A=a$. To illustrate these concepts, suppose that in clinical practice, there is binary treatment $A$ that causes binary outcome $Y$ but their relationship is confounded by binary variable $L$. From probability theory, we can find the mean of $Y$ as:

$$E\left[ Y \right]= \sum_{a,l} E\left[ Y|A=a, L=l \right] P\left[ A=a|L=l \right] P\left[ L=l \right]$$

Now suppose we postulate a hypothetical scenario where we are able to intervene on the treatment received and assign all individuals to receive $A=a$. In this supposed world where we intervene on $A$, we replace the conditional probability mass function $P\left[ A=a|L=l \right]$ with a distribution where all probability is localised to $A=a$, and the probability of any value of *A* other than *a* is zero i.e. an indicator function $1\left( A=a \right)$. Assuming the assumptions of no interference (one participant’s exposure does not affect another participant’s outcome), consistency (one’s potential outcome if treated with $A=a$ is equal to the observed outcome if actually treated with $A=a$), conditional exchangeability (no confounding between $A$ and $Y$ once some covariates are adjusted), and positivity (for every combination of confounders, there is a non-zero probability of being given each treatment option under investigation), we can obtain the mean potential outcome from the observed data with the g-formula:

Eq (A.1)

$$E\left[ Y^{a} \right]= \sum_{a,l} E\left[ Y^{a}|A=a, L=l \right] P\left[ A=a|L=l \right] P\left[ L=l \right]$$

$$= \sum_{l} E\left[ Y^{a}|A=a, L=l \right] P\left[ L=l \right]$$

$$=\sum_{l} E\left[ Y|A=a, L=l \right] P\left[ L=l \right]$$

Since A is binary, we can then formally define an average treatment (causal) effect by taking the difference $E\left[ Y^{1} \right]-E\left[ Y^{0} \right]$.

*The g-formula for a controlled direct effect*

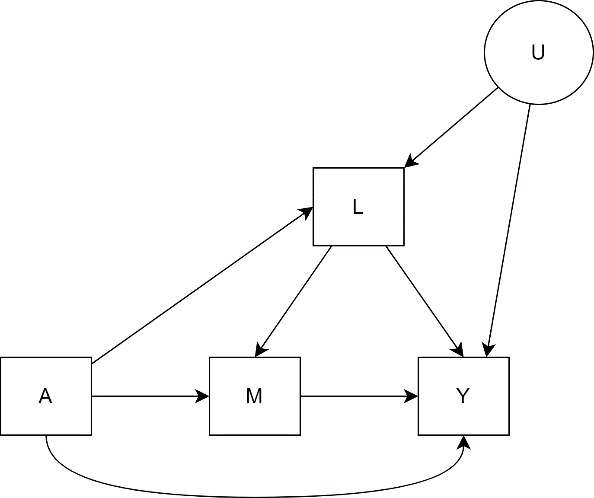

Figure A1: Directed acyclic graph of a point exposure $A$, single mediator $M$, outcome $Y$, mediator-outcome confounder $L$ that is caused by the exposure, and unmeasured variable $U$.

Now suppose we have a setting with baseline, one follow-up visit, and a final visit where the outcome is measured (Figure A1). All variables are binary for simplicity. We make the following suppositions:

I. The data come from a randomised controlled trial and so $A$ represents assigned treatment (treatment vs. placebo).
II. $L$ is measured at the follow-up visit and perhaps represents the presence/absence of some biomarker.
III. $M$, representing adherence, and $Y$, the outcome, are measured at the final visit where we assume $M$ occurs before $Y$.
IV. There is, at least in principle, a conceivable way to intervene on $M$ such that all participants take on the same value of $M$.
V. Biomarker $L$ influences whether a participant subsequently becomes adherent (perhaps due to its role in causing symptoms that discourage adherence) and also influences the outcome $Y.$ Further suppose that the treatment is more likely than placebo to cause changes in the biomarker. This makes $L$ a confounder of the mediator-outcome relationship that is caused by the exposure.
VI. There is some latent variable $U$ that is unmeasured but represents some measure of health that influences values of $L$ and $Y$.

In this setting, we are interested in the controlled direct effect (CDE) of the exposure on Y. That is, all pathways from $A$ to $Y$ that do not go through $M$ because it will be held fixed at the same value for all participants. These pathways are $A \to L\to Y$ and $A\to Y$. Define $Y^{a,m}$ to be the potential outcome if it were possible to intervene at baseline and set exposure to $A=a$ in all participants and intervene during follow-up and be able to ensure that all participants had a desired adherence level $M=m$. The CDE under adherence level $m$ is then defined as:

$${CDE}_{m}=E\left[ Y^{1,m} \right]-E\left[ Y^{0,m} \right]$$

Identification of the CDE from the data requires two assumptions (2):

1. There is no unmeasured confounding in the exposure-outcome relationship (which is satisfied by randomisation); and
2. There is no unmeasured confounding in the mediator-outcome relationship (which is satisfied by controlling for $L$).

Assumption 1 implies that $A⫫U$ and assumption 2 implies $M⫫U | L,A$, where $⫫$ means independence. From these assumptions, $E\left[ Y^{a,m} \right]$ can be identified from the data.

Eq. (A.2)

$$E\left[ Y^{a,m} \right]$$

$$=\sum_{m,l,a,u} E\left[ Y^{a,m}|M=m,L=l, A=a, U=u \right]P(M=m|L=l,A=a,U=u)P\left( L=l | A=a,U=u \right)P(A=a|U=u)P(U=u)$$

$$=\sum_{l,u} E\left[ Y|M=m,L=l, A=a,U=u \right]P\left( L=l | A=a,U=u \right)P\left( U=u \right)$$

$$=\sum_{l,u} E\left[ Y|M=m,L=l, A=a, U=u \right] \frac{P\left( U=u \right)P\left( A=a | U=u \right)P\left( L=l | A=a,U=u \right)}{P\left( A=a | U=u \right)}$$

$$=\sum_{l,u} E\left[ Y|M=m,L=l, A=a, U=u \right] \frac{P\left( L=l,A=a,U=u \right)}{P\left( A=a | U=u \right)}$$

$$=\sum_{l,u} E\left[ Y|M=m,L=l, A=a,U=u \right] \frac{P\left( A=a \right)P\left( L=l | A=a \right)P\left( U=u | L=l,A=a \right)}{P\left( A=a | U=u \right)}$$

$$=\sum_{l,u} E\left[ Y|M=m,L=l, A=a, U=u \right] \frac{P\left( A=a \right)P\left( L=l | A=a \right)P\left( U=u | L=l,A=a \right)}{P\left( A=a \right)}$$

$$=\sum_{l,u} E\left[ Y|M=m,L=l, A=a, U=u \right] \frac{P\left( A=a \right)P\left( L=l | A=a \right)P\left( U=u | M=m,L=l,A=a \right)}{P\left( A=a \right)}$$

$$=\sum_{l} \left( \sum_{u} E\left[ Y|M=m,L=l, A=a, U=u \right]P\left( U=u | M=m,L=l,A=a \right) \right)P\left( L=l | A=a \right)$$

$$=\sum_{l} E\left[ Y|M=m,L=l, A=a \right] P\left( L=l | A=a \right)$$

The third line in equation A.2 for $E\left[ Y^{a,m} \right]$ invokes the consistency assumption, namely $E\left[ Y^{a,m}|M=m, A=a \right]=E\left[ Y|M=m, A=a \right]$, and that intervening on $A$ and $M$ replaces their conditional probability mass functions with the respective indicator functions. Assumptions 1 and 2 to identify the CDE are invoked in lines 7 (see denominator) and 8 (see numerator) of equation A.2 respectively. Though $P\left( A=a | U=u \right)$ is reintroduced in line 4 of equation A.2, we do not sum over values of $A$ since there is only one value.

Scenarios with binary variables could be solved non-parametrically. However, when the covariate set becomes large or contains continuous covariates, the parametric g-formula is needed (also known as g-computation). Essentially, the parametric g-formula uses Monte Carlo simulation to (approximately) solve the high-dimensional sums or integrals but requires parametric models to model the conditional mean of the outcome and conditional probability distributions of covariates like $L$ (3). Each of these parametric models is known as a Q-model.

### **Appendix B: Parametric g-formula steps and modelling assumptions**

**Parametric g-formula steps:**

1. Model the number of GI adverse events (multinomial logistic regression) and the primary outcome (linear regression). See table below for modelling assumptions and specifications.
2. Simulate a sample of $K=10000$ observations by resampling (with replacement) the baseline covariate data. Set a column for arm $A=1$ for all observations.
3. Predict the probabilities of GI adverse events using the model in Step 1 on the dataset with $K$ observations.
4. Obtain Monte Carlo simulations of number of GI adverse events using the predicted probabilities from step 3 on the dataset with $K$ observations.
5. Set a column for adherence $M=1$ for all $K$ observations.
6. Predict the primary outcome using the model in Step 1 for all $K$ observations.
7. Compute the mean of the predicted primary outcome data to obtain an estimate of $E\left[ Y^{A=1,M=1} \right]$.
8. Repeat Steps 2 to 7 but amending $A=0$ in Step 2 to obtain estimate of $E\left[ Y^{A=0,M=1} \right]$.
9. Calculate the point estimate of the CDE, $E\left[ Y^{A=1,M=1} \right]-E\left[ Y^{A=0,M=1} \right]$.
10. Repeat Steps 1 to 9 using $B=5000$ bootstrap replicates of the original data to derive a bootstrap (either percentile-based or BCa) 95% confidence interval of the CDE. The standard deviation of bootstrap estimates is also calculated as ${SE}_{boot}$ for use in the probabilistic bias analysis (Appendix C). The relatively high value of $B$ was chosen to balance computation time and stabilisation of the BCa interval.

**Table of modelling assumptions**

All explanatory variables have an interaction term with treatment arm in all models. Number of GI adverse events is treated as a categorical variable (reference category: 0 events) when included as a predictor. The quadratic and cubic terms for continuous covariates in the primary outcome regression model were decided on after an initial natural course analysis with only linear terms; we observed a qualitatively closer agreement between the eCDFs when non-linear terms were included. The quadratic and cubic terms for continuous covariates were not included in the models for GI events or adherence due to warnings suggesting separation. A summary is displayed in Table B1 below.

| **Variable** | **Parametric distribution** | **Explanatory variables** | **Modelling approach** |
| --- | --- | --- | --- |
| Number of GI adverse events (0, 1, ≥2 events) | Multinomial | Treatment arm, age, sex, obesity, history of depression, NEADL, polypharmacy, baseline 4-metre walk speed | Multinomial logistic regression with the multinom function in the *nnet* package (4). Age, NEADL, and baseline 4-metre walk speed were standardised to the observed sample mean and standard deviation. |
| 4-metre walk speed at four months | Normal | Treatment arm, age, sex, obesity, history of depression, NEADL, polypharmacy, baseline 4-metre walk speed, number of GI adverse events, adherence | Linear regression. Age, NEADL, and baseline 4-metre walk speed were standardised to the observed sample mean and standard deviation. Linear, quadratic, and cubic terms were included for age, PCS, and baseline 4-metre walk speed. |
| Adherence | Binomial | Treatment arm, age, sex, obesity, history of depression, NEADL, polypharmacy, baseline 4-metre walk speed, number of GI adverse events | Logistic regression. Age, NEADL, and baseline 4-metre walk speed were standardised to the observed sample mean and standard deviation. Adherence is only modelled for the natural course analysis. |

Table B1: Summary of modelling specifications. NEADL, Nottingham Extended Activities of Daily Living; GI gastrointestinal.

### **Appendix C: Probabilistic bias analysis (PBA)**

The extent of misclassification is defined by two bias parameters: the false positive rate (FPR) i.e., the proportion of participants who are observed as adherent (based on returned tablet counts) but were truly non-adherent (based on actual consumption); and the false negative rate (FNR) i.e., the proportion of participants who are observed as non-adherent (based on returned tablet counts) but were truly adherent (based on actual consumption). For our PBA, the FNR is fixed to 0 as false negatives were judged to be highly implausible; it implies that participants consumed tablets then replaced what they had consumed to appear as though they had not.

Briefly, the process of PBA involves first drawing a value of the FPR from a Beta distribution representing our beliefs about what the FPR value is. The literature refers to this as a bias parameter distribution, but it may be convenient to call this a ‘prior’ representing our beliefs about the FPR. This has advantages over choosing a single value of the FPR since we can never be certain of the actual FPR, so it is more reasonable to entertain a range of plausible FPR values, some of which are more likely than others. Then, we (Monte Carlo) simulate the “true adherence” indicator for each individual using this drawn FPR value alongside the observed data, before repeating the parametric g-formula analyses with this “true adherence” variable. This entire process is repeated with a large number of iterations to obtain bias-adjusted estimates of the CDE. Details on PBA are described below but further information are available elsewhere (5).

We first conducted a literature review to gather information on plausible values of the FPR based on papers comparing pill count-and Medication Event Monitoring System (MEMS) based adherence. We chose MEMS as the referent as it is considered the closest to a gold standard for medication adherence (6). One study author (SH) retrieved 65 results from PubMed on 18 March 2024 using the search terms: (adherence or compliance) and "event monitoring" and ("pill count" or "medication possession ratio"). Studies were included if: 1) an adherence threshold was (or could be) defined; 2) MEMS- vs. pill count- (or MPR) based adherence were compared; and 3) enough data were reported to obtain or calculate an estimate of the FPR. Only full original articles were included, but reference sections of evidence synthesis papers or opinion/commentary papers were checked. Studies conducted in children and adolescents were excluded. Additional articles were obtained through Google Scholar searches. A line listing of eight included papers is presented as Appendix Table C1.

We constructed four FPR priors to represent four scenarios. For scenario 1, we used a $\mathrm{Beta}\left( 33.3, 231 \right)$ by requiring that 0.049, 0.125, and 0.161 be the 5^th^, 50^th^, and 95^th^ percentiles respectively (Figure C1). This scenario represents a very high certainty in our beliefs that there is a low degree of misclassification.

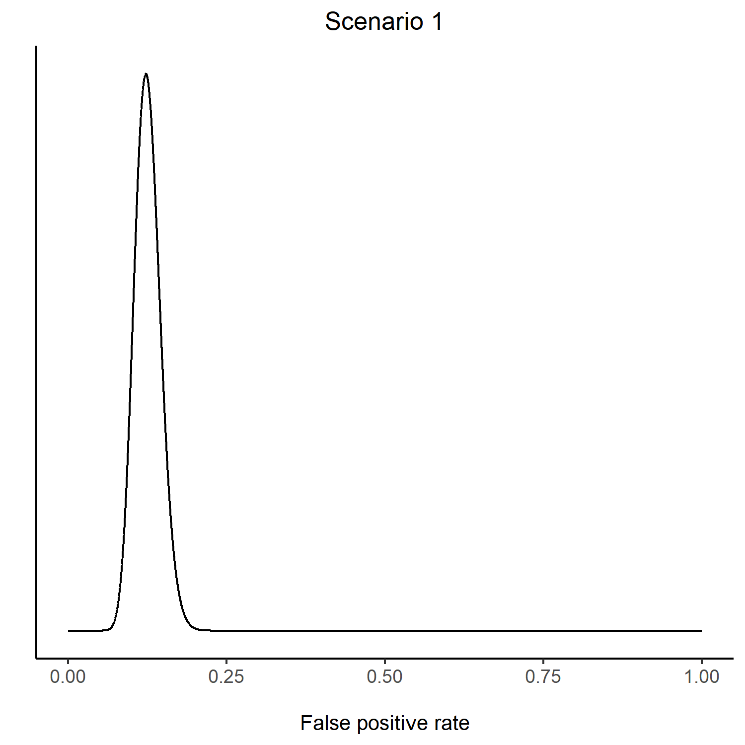

Figure C1: Illustration of the scenario 1 prior

For scenario 2, we used a $\mathrm{Beta}\left( 355, 205 \right)$ by requiring that 0.478, 0.634, and 0.667 be the 5^th^, 50^th^, and 95^th^ percentiles respectively (Figure C2). This scenario represents a very high certainty in our beliefs that there is a severe degree of misclassification.

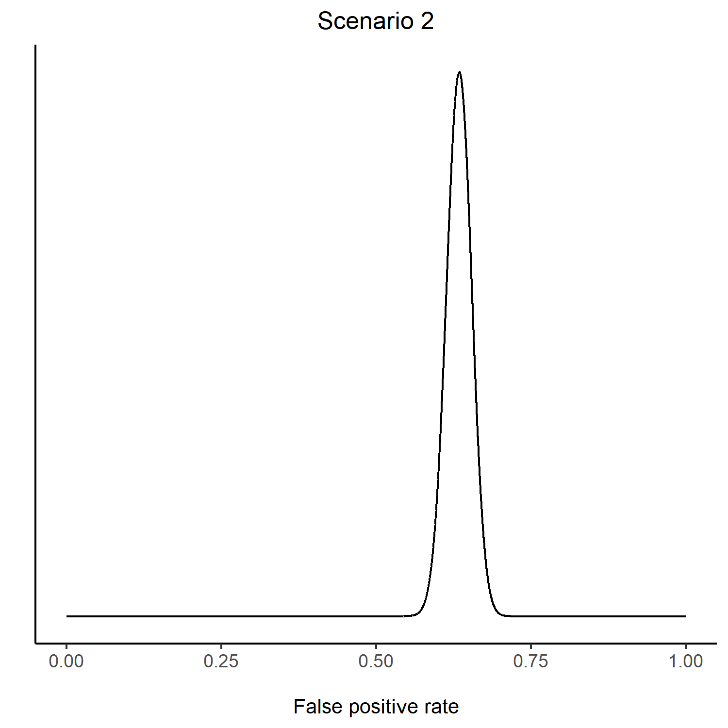

Figure C2: Illustration of the scenario 2 prior

For scenario 3, we used a $\mathrm{Beta}\left( 4, 26 \right)$ by requiring that 0.049, 0.125, and 0.40 be the 5^th^, 50^th^, and 95^th^ percentiles respectively (Figure C3). This scenario represents more moderate certainty in our beliefs that there is a low degree of misclassification.

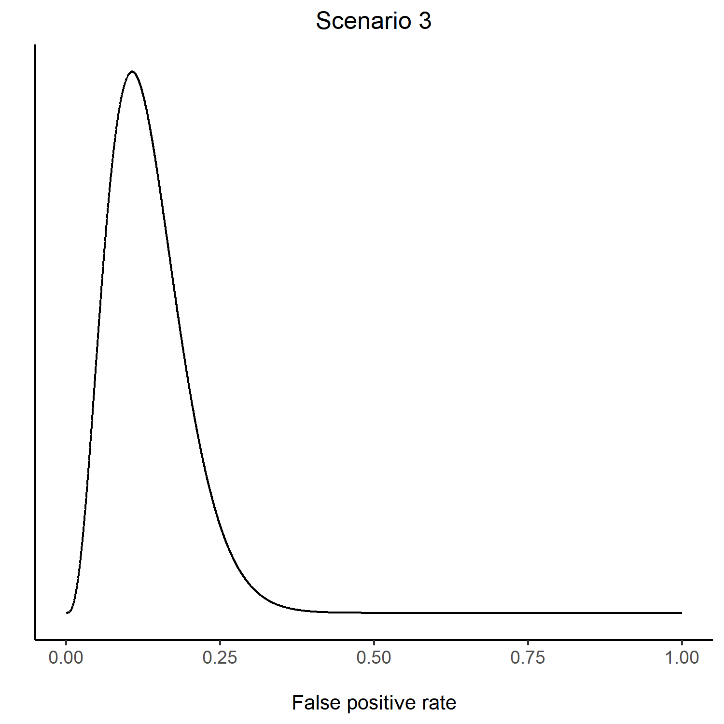

Figure C3: Illustration of the scenario 3 prior

For scenario 4, we used a $\mathrm{Beta}\left( 39.8, 25 \right)$ by requiring that 0.400, 0.615, and 0.710 be the 5^th^, 50^th^, and 95^th^ percentiles respectively (Figure C4). This scenario represents more moderate certainty in our beliefs that there is a severe degree of misclassification.

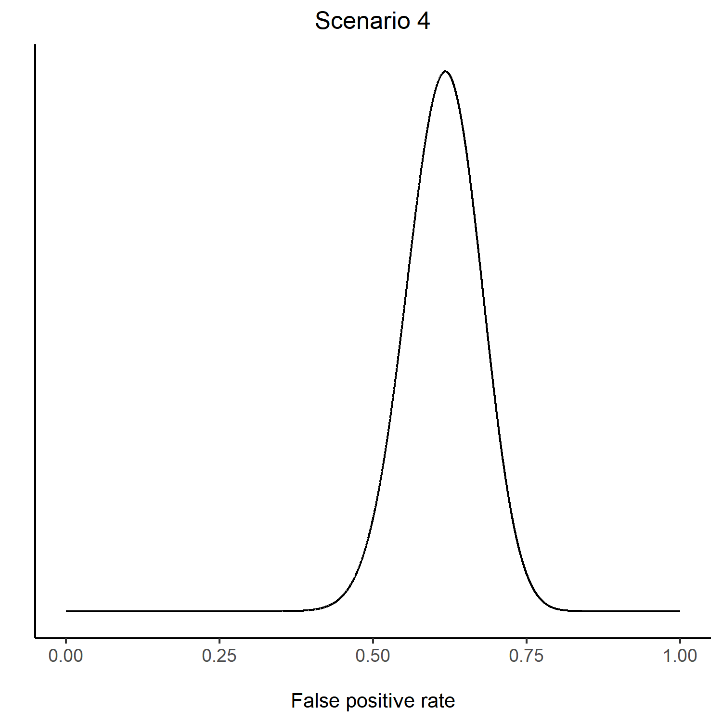

Figure C4: Illustration of the scenario 4 prior

These distributions for scenarios 1 and 2 have approximately the same standard deviation of 0.02, and scenarios 3 and 4 have approximately the same standard deviation of 0.06. We assumed non-differential misclassification between arms i.e., the extent of misclassification is the same in both arms.

Using a randomly drawn FPR value and an FNR of 0, we can calculate the probability of true adherence in each arm, followed by the probability of being truly adherent given observed adherence (positive predictive value, PPV) and the probability of being truly non-adherent given observed non-adherence (negative predictive value, NPV). A “true adherence” column was then constructed by simulating from a Bernoulli distribution whose parameter was the PPV for intervention participants and NPV for control participants (7). The parametric g-formula was then run (without the bootstrapping step) but using the new “true adherence” variable as the mediator. A random normal deviate $Z \sim N(0,1)$ multiplied by ${SE}_{boot}=0.38$ (see Appendix B) is then subtracted from each estimate to reintroduce random error into the estimates. This entire process is repeated with 5,000 FPR samples to obtain bias-adjusted estimates of the CDE. The median of the iterations is used as the point estimate of the misclassification bias-adjusted CDE with the percentile-based 95% simulation interval, representing our uncertainty around this point estimate that incorporates both systematic (due to misclassification) and random error.

| **Author** | **Patient group and treatment** | **Study type** | **Sample size** | **Mean age (SD)** | **Adherence threshold** | **Timeframe** | **Reference standard** | **Prevalence of adherence by MEMS** | **Prevalence of adherence by pill count** | **FPR** |
| --- | --- | --- | --- | --- | --- | --- | --- | --- | --- | --- |
| Bangsberg 2001 * (8) | HIV and ART | Prospective cohort | 42 | - | ≥80% | 3 months | MEMS | 42.9% | 42.9% | 0.125 |
| Brain 2014 † (9) | Schizophrenia and antipsychotics | Prospective cohort | 117 | 46 | ≥80% | 12 months | MEMS | 73% | 71% | 0.161 |
| Jasti 2006 (10) | Pregnant women and iron supplements | RCT | 51 | 23 | ≥75% | 6.5-7 months | MEMS | 54.9% | 72.5% | 0.478 |
| Márquez-Contreras 2018 (11) | HTN and antihypertensives | Prospective cohort | 102 | 61.1 (9.1) | ≥80% | 6 months | MEMS | - | - | 0.049 |
| Ruslami 2008 (12) | TB and TB treatment + pyridoxine | Prospective cohort | 30 | 32 | 100% | 1 month | MEMS | 57% | 53% | 0.400 |
| Van den Boogaard 2011 (13) | TB and TB treatment | Prospective cohort | 50 | 41.6 (14) | ≥95% | 4 months | MEMS | 79% | - | 0.710 |
| van Onzenoort 2010 (14) | Mild-moderate HTN and Lisinopril + hydrochlorothiazide | RCT | 228 | 57 (10) | ≥90% | 1 year | MEMS | 60.5% | 71.9% | 0.634 |
| Winkler 2002 (15) | Type 2 diabetes and sulfonylureas | Prospective cohort | 19 | 68.8 (10.7) | ≥80% | 2 months | MEMS | 84.2% | 89.5% | 0.667 |

Table C1: Studies comparing pill count-based adherence and MEMS-based adherence*.* ART, antiretroviral therapy; FPR, false positive rate; HIV, human immunodeficiency virus; HTN, hypertension; RCT, randomised controlled trial; TB, tuberculosis. Adherence based on MEMS was the reference standard.

* The “adjusted” electronic monitored dose adherence measurements were used.

† Values for FPR were reconstructed from the concordance statistic and the various marginal probabilities of adherence.

### **Appendix D: The MET-PREVENT study group members and affiliations**

Andrew P Clegg^1^

Helen Hancock^2^

Claire McDonald^3,4^

Avan A Sayer^3,4^

Claire J Steves^5,6^

Thomas von Zglinicki^7^

1. Academic Unit for Ageing & Stroke Research, University of Leeds, Bradford Teaching Hospitals NHS Foundation Trust, Bradford, UK

2. Newcastle Clinical Trials Unit, Newcastle University, Newcastle upon Tyne, UK

3. AGE Research Group, Translational and Clinical Research Institute, Faculty of Medical Sciences, Newcastle University, Newcastle upon Tyne, UK

4. NIHR Newcastle Biomedical Research Centre, Newcastle upon Tyne Hospitals NHS Foundation Trust, Cumbria, Northumberland, Tyne and Wear NHS Foundation Trust and Newcastle University, Newcastle upon Tyne, UK

5. Department of Twin Research and Genetic Epidemiology, School of Lifecourse Sciences and Population Health, London UK

6. Department of Ageing and Health, Guys and St Thomas’ NHS Foundation Trust, London UK

7. Ageing Biology Labs, Newcastle University Biosciences Institute, Newcastle University, Newcastle upon Tyne, UK
